## Supplemental material for "Cross-GWAS coherence test at the gene and pathway level"

July 2022

Daniel Krefl[a,b] and Sven Bergmann[a,b,c]
[a] *Department of Computational Biology, University of Lausanne, Switzerland*
[b] *Swiss Institute of Bioinformatics, Lausanne, Switzerland*
[c] *Dept. of Integrative Biomedical Sciences, University of Cape Town, Cape Town, South Africa*

### I. GENERAL PRODUCT-NORMAL

Let us briefly consider the distribution for $xy$ with $x \sim \mathcal{N}(\mu_x, \sigma_x^2)$ and $y \sim \mathcal{N}(\mu_y, \sigma_y^2)$, *i.e.*, the product-normal for non-standardized Gaussian random variables. The moment generating function reads [1]

$$M_{x,y}(\nu) = \frac{e^{\frac{(\mu_x^2\sigma_y^2+\mu_y^2\sigma_x^2-2\varrho\mu_x\mu_y\sigma_x\sigma_y)\nu^2+2\mu_x\mu_y\nu}{2(1-\sigma_x\sigma_y(1-\varrho)\nu)(1+\sigma_x\sigma_y(1+\varrho)\nu)}}}{\sqrt{(1-\sigma_x\sigma_y(1+\varrho)\nu)(1+\sigma_x\sigma_y(1-\varrho)\nu)}} \,. \tag{1}$$

Note that for, either, $\mu_x = 0$ or $\mu_y = 0$, the factorization of the main text still holds. Defining $\kappa_x = \frac{\mu_x}{\sigma_x}$ and $\kappa_y = \frac{\mu_y}{\sigma_y}$, the exponent of the exponential in $M_{x,y}\left(\frac{2\nu}{\sigma_x\sigma_y}\right)$ can be split for the general non-correlated case ($\varrho = 0$) into

$$\frac{(\kappa_x^2+\kappa_y^2+2\kappa_x\kappa_y)\nu}{2(1-2\nu)} - \frac{(\kappa_x^2+\kappa_y^2-2\kappa_x\kappa_y)\nu}{2(1+2\nu)} \,.$$

We deduce that we can still factorize the moment generating function, such that

$$\begin{aligned} xy \sim & \frac{\sigma_x\sigma_y}{2}\left[\chi_1^2\left(\frac{1}{2}(\kappa_x^2+\kappa_y^2+2\kappa_x\kappa_y)\right)\right] \\ & - \frac{\sigma_x\sigma_y}{2}\left[\chi_1^2\left(\frac{1}{2}(\kappa_x^2+\kappa_y^2-2\kappa_x\kappa_y)\right)\right] , \end{aligned} \tag{2}$$

with $\chi_1^2(c)$ the non-central $\chi^2$ distribution with one degree of freedom. Hence, the general product-normal can be expressed as a linear combination of non-central $\chi_1^2$ distributions, for $\varrho = 0$.

### II. PROBABILITY DENSITY FUNCTIONS

*a. Product-Normal:* Making use of the relation (2) for standardized variables and expressed in terms of gamma distributions, the pdf, denoted as $f$, of the product-normal distribution can be calculated analytically via convolution (*cf.*, [2])

$$f_{\xi-\zeta}(x) = \frac{e^{\frac{x}{1-\varrho}}}{\pi\sqrt{1-\varrho^2}} \int_{\max(0,x)}^{\infty} dy (y^2-xy)^{-1/2} e^{-\frac{2y}{1-\varrho^2}} \,.$$

| $\varrho$ | 0 | 0.3 | 0.6 | 0.9 |
|---|---|---|---|---|
| MSE | $3.3\times10^{-15}$ | $3.5\times10^{-15}$ | $4.6\times10^{-15}$ | $2.1\times10^{-12}$ |

TABLE I: Mean squared error (MSE) between numerical integration of (3) and Davies algorithm for various $\varrho$ and 1000 arguments evenly spaced in the range $[-4, 4]$.

Completing the square and invoking a hyperbolic substitution, we arrive at

$$\begin{aligned} f_{\xi-\zeta}(x) &= \frac{e^{\frac{\varrho x}{1-\varrho^2}}}{\pi\sqrt{1-\varrho^2}} \int_0^{\infty} dt\, e^{-\frac{|x|}{1-\varrho^2}\cosh(t)} \\ &= \frac{e^{\frac{\varrho x}{1-\varrho^2}}}{\pi\sqrt{1-\varrho^2}} K_0\left(\frac{|x|}{1-\varrho^2}\right) , \end{aligned} \tag{3}$$

with $K_0$ the modified Bessel function of the second kind at zero order. The result above for $f$ agrees with the previous derivations of [3, 4]. Note that the analytic calculation of the corresponding cdf requires the solution of an integral of the type $\int_x^{\infty} dt\, e^{at}K_0(t)$ . We are not aware of a known closed-form solution.

For illustration, we plot the pdf for $\varrho = 1/2$ together with the corresponding histogram sampled from (2) expressed as a difference of gamma distributions in figure 1. We also show the cdf obtained via numerical integration of (3). We verified that the numerical integration matches the results obtained via Davies' algorithm for the cdf calculation for various $\varrho$, *cf.*, table I.

Note that since the pdf of the non-central $\chi^2$ distribution includes a Bessel function, analytic calculation of the pdf of $xy$ in the more general case of section I is more complicated than in (3), and will not be discussed here.

*b. Variance-Gamma:* Consider a random variable $X$ distributed according to

$$X \sim [\Gamma(h/2, g)] - [\Gamma(h/2, g)] =: VG(h, g) \,. \tag{4}$$

The corresponding pdf can be calculated similarly as above via convolution. We infer

$$f_{\xi-\zeta}(x) = \frac{e^{\frac{x}{g}}}{\Gamma(h/2)^2 g^h} \int_{\max(0,x)}^{\infty} dy\, (y^2-xy)^{h/2-1} e^{-\frac{2y}{g}} \,.$$

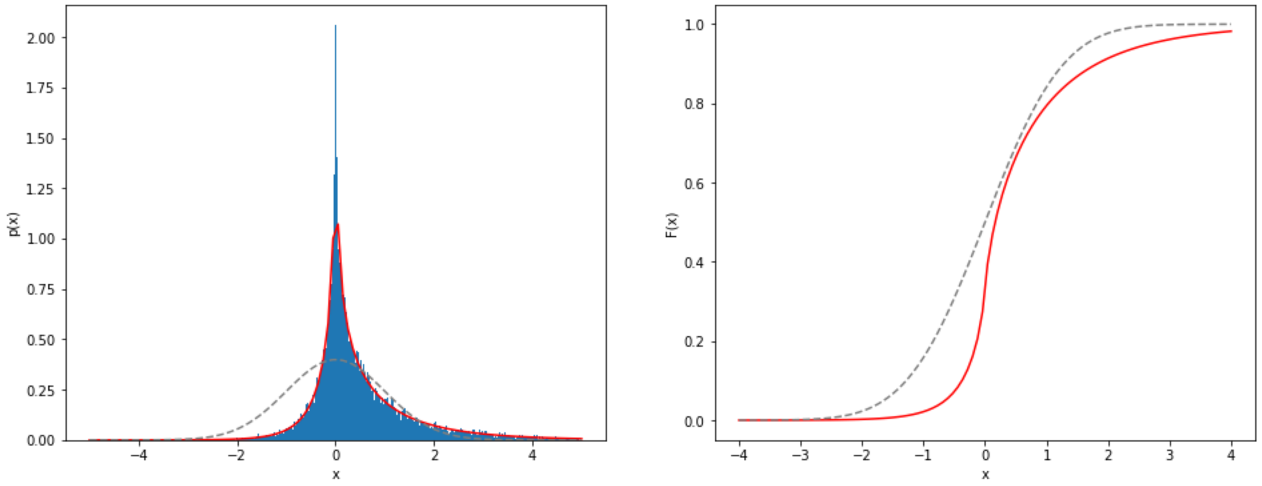


FIG. 1: Left: Histogram of the dependent product normal distribution with $\rho = 0.5$ obtained via subtracting 50.000 pairs of random samples of difference of gamma distributions. The red line marks the pdf given in (3). Right: The corresponding cdf obtained via Davies algorithm. For comparison, the dashed gray lines show the corresponding normal quantities.

Completing the square and using as before hyperbolic substitution, we arrive at

$$f_{\xi-\zeta}(x) = \frac{x^{h-1}}{2^{h-1}\Gamma(h/2)^2 g^h}\int_0^\infty dt\,\sinh(t)^{h-1}e^{-\frac{|x|}{g}\cosh(t)} = \frac{K_{\frac{h-1}{2}}(|x|/g)}{g\sqrt{\pi}\,\Gamma(h/2)}\left(\frac{|x|}{2g}\right)^{\frac{h-1}{2}}, \tag{5}$$

with $K_n$ a modified Bessel function of second kind at order $n$. Using the integral $\int_0^\infty dt\, t^{\mu-1}K_\nu(t) = 2^{\mu-2}\Gamma(\frac{\mu-\nu}{2})\Gamma(\frac{\mu+\nu}{2})$, we easily verify that $\int_0^\infty dx\, f_{\xi-\zeta}(x) = \frac{1}{2}$. Hence, due to symmetry the pdf is well normalized. However, we are not aware of closed-form solutions for Bessel function integrals of the type $\int_x^\infty dt\, t^\nu K_\nu(t)$, which are needed to provide a closed-form expression for the cdf.

The distribution given by (5) occurred before in the finance domain as a special case of the *variance-gamma* distribution [5]. It can be traced back further to the distribution of the bivariate correlation. In detail, the gamma-variance distribution corresponds to the off-diagonal marginal of a two-dimensional Wishart distribution, which models the covariance matrix [6]. However, to the best of our knowledge, what is new is the expression in terms of the difference distribution in equation (4).

For $h = n$ and $g = 1$, we have

$$X \sim VG(n,1) = \frac{1}{2}[\chi^2_n] - \frac{1}{2}[\chi^2_n]\,. \tag{6}$$

Hence, for $n = 1$, we obtain the product-normal distribution with $\varrho = 0$ as discussed in the previous section. For general $n$, we can view $VG(n,1)$ as the distribution of a sum of $n$ independently distributed product-normal random variables. In particular, we can use Davies' algorithm to calculate the cdf for $VG(n,1)$ exactly at the desired precision.

### III. SIMULATIONS: $N = 2$

The importance of correcting for the inter-dependence between the elements of $w$ and $z$ in the index $I = \sum_i w_i z_i$ introduced in the main text can be seen easily in the $N = 2$ case. Consider the covariance matrices,

$$\Sigma = \Sigma_w = \Sigma_z = \begin{pmatrix} 1 & r \\ r & 1 \end{pmatrix},$$

such that $w \sim \mathcal{N}(0,\Sigma)$ and $z \sim \mathcal{N}(0,\Sigma)$, and with $r$ varying. In figure 2 we show various significance threshold curves of $I$ for varying $r$, as calculated from the to $I$ corresponding distribution expressed as a linear combination of $\chi^2$ distributions, *cf.*, the main text. Clearly, for increasing inter-element correlation, the significance threshold level rises. The magnitude of the effect increases with the desired level of significance.

### IV. ACCOUNTING FOR SAMPLE OVERLAP

For GWAS, the populations used to investigate two traits for co-significant signals might overlap. In particular, it may even be that the GWAS for the two traits have been conducted for the same population (full overlap). If

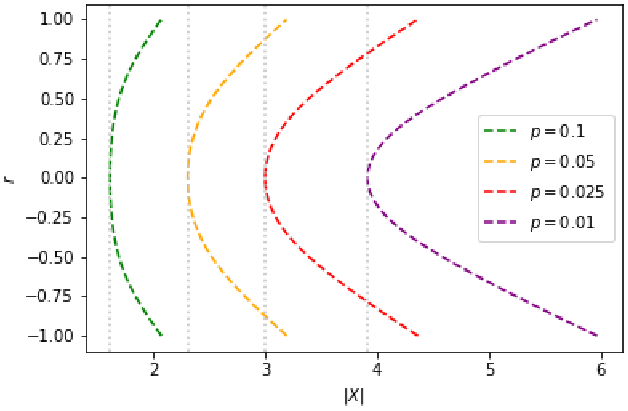


FIG. 2: Significance threshold curves of $I$ in the two-element case under variation of the inter-element correlation ($y$-axis). The $x$-axis corresponds to the argument of the tail probability $p = 1 - F_I(X)$, with $F_I$ the cdf of the distribution of $I$. The gray dotted lines mark the minimal value obtained for zero correlation.

there is in addition a strong phenotypic correlation between the traits, a danger for a significant biased result is present, as long as one does not correct for this effect induced by the sample overlap.

Let us consider for simplicity first the case of full population sample overlap. For same population (and so genotype matrix $X$), and for two different traits $y^{(1)}$ and $y^{(2)}$, we have as multi-variate models

$$\begin{aligned} y^{(1)} &= X\alpha^{(1)} + \epsilon^{(1)}\,, \\ y^{(2)} &= X\alpha^{(2)} + \epsilon^{(2)}\,, \end{aligned} \tag{7}$$

with true effect sizes $\alpha^{(i)}$ and residuals $\epsilon^{(i)} \sim \mathcal{N}(0, \mathbb{1})$.

Our test null assumption is that $\alpha^{(i)} = 0$. Therefore,

$$\mathrm{cov}(y^{(1)}, y^{(2)}) = \mathrm{cov}(\epsilon^{(1)}, \epsilon^{(2)}) = (\epsilon^{(1)})^T \epsilon^{(2)} := \zeta\,.$$

We can assume that the residuals are independent for independent populations but generally $\zeta \neq 0$ for overlapping populations. This implies

$$\begin{pmatrix} \epsilon^{(1)} \\ \epsilon^{(2)} \end{pmatrix} \sim \mathcal{N}\left(0, \begin{pmatrix} \mathbb{1} & \zeta\mathbb{1} \\ \zeta\mathbb{1} & \mathbb{1} \end{pmatrix}\right)\,. \tag{8}$$

Recall from the main text that the z-scored null effect estimates read (as vector over some region)

$$z = \frac{1}{\sqrt{n}} x^T \epsilon \sim \mathcal{N}(0, \Sigma),$$

with $\Sigma := \frac{1}{n} x^T x$ and $x_i$ columns of $X$. It follows that

$$\begin{pmatrix} z^{(1)} \\ z^{(2)} \end{pmatrix} = \frac{1}{\sqrt{n}} \begin{pmatrix} x^T & \\ & x^T \end{pmatrix} \begin{pmatrix} \epsilon^{(1)} \\ \epsilon^{(2)} \end{pmatrix} \sim \mathcal{N}\left(0, \begin{pmatrix} \Sigma & \zeta\Sigma \\ \zeta\Sigma & \Sigma \end{pmatrix}\right)\,. \tag{9}$$

Using block Gaussian elimination, one can show that

$$\begin{pmatrix} \Sigma & \zeta\Sigma \\ \zeta\Sigma & \Sigma \end{pmatrix} = \begin{pmatrix} \mathbb{1} & \\ \zeta\mathbb{1} & \mathbb{1} \end{pmatrix} \begin{pmatrix} \Sigma & \\ & (1-\zeta^2)\Sigma \end{pmatrix} \begin{pmatrix} \mathbb{1} & \zeta\mathbb{1} \\ & \mathbb{1} \end{pmatrix}\,. \tag{10}$$

Hence,

$$\begin{pmatrix} z^{(1)} \\ z^{(2)} \end{pmatrix} \sim \begin{pmatrix} \mathbb{1} & \\ \zeta\mathbb{1} & \mathbb{1} \end{pmatrix} \begin{pmatrix} x \\ y \end{pmatrix} = \begin{pmatrix} x \\ y + \zeta x \end{pmatrix}\,, \tag{11}$$

with

$$\begin{pmatrix} x \\ y \end{pmatrix} \sim \mathcal{N}\left(0, \begin{pmatrix} \Sigma & \\ & (1-\zeta^2)\Sigma \end{pmatrix}\right)\,. \tag{12}$$

We conclude that

$$\mathrm{cov}(z_i^{(1)}, z_j^{(2)}) = \mathrm{cov}(x_i, y_j + \zeta x_j) = \zeta\,\mathrm{cov}(x_i, x_j) = \zeta\Sigma_{ij}\,.$$

Consider now a matrix $U$ such that $\Sigma = U\Lambda U^T$ with $\Lambda$ the diagonal matrix of eigenvalues. Clearly, $UU^T = 1$. We then have

$$\begin{aligned} z^{(1)} &= U^T \sqrt{\Lambda} w \\ z^{(2)} &= U^T \sqrt{\Lambda} v \end{aligned} \tag{13}$$

with $w, v \sim \mathcal{N}(0, \mathbb{1})$. The correlation between the new variables $w$ and $v$ can be calculated to be given by

$$\begin{aligned} \mathrm{corr}(w_i, v_j) &= \frac{\mathrm{cov}(w_i, v_j)}{\sqrt{\mathrm{var}(w_i)}\sqrt{\mathrm{var}(v_j)}} \\ &= \frac{\mathrm{cov}(\frac{1}{\sqrt{\lambda_i}}(Uz^{(1)})_i, \frac{1}{\sqrt{\lambda_j}}(Uz^{(2)})_j)}{\sqrt{\mathrm{var}(\frac{1}{\sqrt{\lambda_i}}(Uz^{(1)})_i)}\sqrt{\mathrm{var}(\frac{1}{\sqrt{\lambda_j}}(Uz^{(2)})_j)}} \\ &= \frac{\sum_{k,l} U_{ik}^T U_{lj} \mathrm{cov}(z_k^{(1)}, z_l^{(2)})}{\sqrt{\mathrm{var}((Uz^{(1)})_i)}\sqrt{\mathrm{var}((Uz^{(2)})_j)}} \\ &= \frac{\sum_{k,l} U_{ik}^T U_{li} \mathrm{cov}(z_k^{(1)}, z_l^{(2)})}{\sqrt{\lambda_i \lambda_j}} \\ &= \zeta \frac{\sum_{k,l} U_{ik}^T \Sigma_{kl} U_{lj}}{\sqrt{\lambda_i \lambda_j}} \\ &= \zeta \frac{\Lambda_{ij}}{\sqrt{\lambda_i \lambda_j}}\,. \end{aligned} \tag{14}$$

It follows that

$$\mathrm{corr}(w_i, v_i) = \begin{cases} \zeta & i = j \\ 0 & i \neq j \end{cases}\,. \tag{15}$$

*a. Coherence test* Recall from the main text (Methods, *Product-Normal distribution*) that the product distribution for correlated $\mathcal{N}(0, 1)$ variables with coefficient $\varrho$ can be expressed as

$$w_i v_i \sim \frac{1+\varrho}{2}[\chi_1^2] - \frac{1-\varrho}{2}[\chi_1^2]\,.$$

We conclude that for the corrected null distribution with identical populations for the two traits

$$(z^{(1)})^T z^{(2)} \sim \sum_i \lambda_i \left(\frac{1+\zeta}{2}\right)[\chi_1^2] - \sum_i \lambda_i \left(\frac{1-\zeta}{2}\right)[\chi_1^2]. \tag{16}$$

The coefficient $\zeta$ is given by the phenotypic correlation. As one would expect, for identical traits ($\zeta = 1$), the above turns into the standard $\chi^2$ test of Pascal [8]. For $\zeta = 0$, we end up with the formula (2.2) of the main text.

The generalization to partial sample overlap is straightforward. We can assume that the samples are sorted such that the overlapping samples form the first $n_o$ rows, followed by the $n_i^{(x)}$ independent samples with $n_o + n_i^{(1)} + n_i^{(2)} = n$. Correspondingly, the residuals read

$$\begin{pmatrix} \epsilon^{(1)} \\ \epsilon^{(2)} \end{pmatrix} \sim \mathcal{N}\left(0, \begin{pmatrix} \mathbb{1}_{n_o\times n_o} & & \zeta\mathbb{1}_{n_o\times n_o} & \\ & \mathbb{1}_{n_i^{(1)}\times n_i^{(1)}} & & \\ \zeta\mathbb{1}_{n_o\times n_o} & & \mathbb{1}_{n_o\times n_o} & \\ & & & \mathbb{1}_{n_i^{(2)}\times n_i^{(2)}} \end{pmatrix}\right), \tag{17}$$

and the z-scored null effect estimates

$$\begin{pmatrix} z^{(1)} \\ z^{(2)} \end{pmatrix} = \begin{pmatrix} \frac{1}{\sqrt{n_o+n_i^{(1)}}}(x^{(1)})^T & \\ & \frac{1}{\sqrt{n_o+n_i^{(2)}}}(x^{(2)})^T \end{pmatrix} \begin{pmatrix} \epsilon^{(1)} \\ \epsilon^{(2)} \end{pmatrix}, \tag{18}$$

with the first $n_o$ rows of $x^{(1)}$ identical to $x^{(2)}$.

We assume that the population covariance matrix $\Sigma$ is well approximated by the sub-populations of both GWAS, *i.e.*,

$$\frac{1}{n_o + n_i^{(k)}}(x^{(k)})^T\, \mathbb{1}_{(n_o+n_i^{(k)})\times(n_o+n_i^{(k)})}\, x^{(k)} \approx \Sigma\,.$$

For the off-diagonal parts, we have

$$\frac{\left(x^{(k)}\right)^T \begin{pmatrix} \mathbb{1}_{n_o\times n_o} & \\ & \end{pmatrix} \left(x^{(l)}\right)}{\sqrt{n_i^{(1)}+n_o}\sqrt{n_i^{(2)}+n_o}} \approx \frac{\Sigma}{\sqrt{1+\frac{n_i^{(1)}}{n_o}}\sqrt{1+\frac{n_i^{(2)}}{n_o}}},$$

under the assumption that the overlapping sub-population also approximates the population covariance structure well.

It follows that the sample overlap calculation proceeds as for the full overlap case discussed above, but with

$$\zeta \to \frac{n_o}{\sqrt{n^{(1)}}\sqrt{n^{(2)}}}\,\zeta\,, \tag{19}$$

with $n^{(k)} = n_o + n_i^{(k)}$ the individual GWAS sample sizes. Note that for $n_o$ sufficiently small the correction factor vanishes, as it should be.

The calculation of $\zeta$ requires knowledge of the phenotypic correlation. However, often only public GWAS summary statistics are at hand. Fortunately, in this case, one can use LD score regression [9] to obtain an estimation, as the intercept term thereof precisely corresponds to the correction factor (19).

In order to illustrate the impact of such a non-zero correction factor, we consider a multivariate normal distribution with correlation matrix $\Sigma$ of dimension 100. $\Sigma$ is taken to be glued together from two 50-dimensional correlation matrices with off-diagonal elements set identical to 0.2 and two off-diagonal block matrices with elements of 0.2 multiplied by various $\zeta$. This setup simulates (9).

We calculate $p$-values for the index $(z^{(1)})^T z^{(2)}$ given in equation (16) for 1000 random samples with and without correction factor $\zeta$. Corresponding QQ-plots are shown in figure 3 and 4. We observe that inclusion of $\zeta$ indeed removes the bias.

*b. Ratio test* The index of the ratio test reads (see Results, section *Ratio test* of the main text)

$$R = \frac{(z^{(1)})^T z^{(2)}}{(z^{(1)})^T z^{(1)}}, \tag{20}$$

with $z^{(i)}$ defined as above. (Similarly for interchanged indices.) We have

$$\begin{aligned} \Pr(R \le r) &= \Pr\left((z^{(1)})^T z^{(2)} \le r (z^{(1)})^T z^{(1)}\right) \\ &= \Pr\left((z^{(1)})^T (z^{(2)} - r\, z^{(1)}) \le 0\,.\right) \end{aligned} \tag{21}$$

We can linearly transform variables such that

$$\Pr(R \le r) = \Pr(w^T \Lambda (v - r\, w) \le 0)\,.$$

Under the re-definition $u := v - r\,w$ we arrive at

$$\Pr(R \le r) = \Pr(w^T \Lambda u \le 0) = F_{\bar{w}\bar{u}}(0)\,,$$

as in the main text. However, because of the non-vanishing correlation (15), we now have instead $\bar{w} \sim \mathcal{N}(0, \Lambda)$ and $\bar{u} \sim \mathcal{N}(0, (1 + r^2 - 2\zeta r)\Lambda)$. In particular,

$$\text{cov}(w_i, u_j) = \text{cov}(w_i, v_j) - r\delta_{ij} = (\zeta - r)\delta_{ij}\,.$$

With $\text{Var}(w_i) = 1$ and $\text{Var}(u_j) = (1 + r^2 - 2\zeta r)$ we obtain for the corresponding correlation

$$\text{corr}(w_i, u_j) = \frac{(\zeta - r)\delta_{ij}}{\sqrt{(1 + r^2 - 2\zeta r)}}\,.$$

Therefore, we deduce similar as in the main text that

$$\begin{aligned} \bar{w}\bar{u} \sim \sum_i & \frac{\lambda_i\sqrt{1 + r^2 - 2\zeta r}(1+t)}{2}[\chi_1^2] \\ & - \frac{\lambda_i\sqrt{1 + r^2 - 2\zeta r}(1-t)}{2}[\chi_1^2]\,, \end{aligned} \tag{22}$$

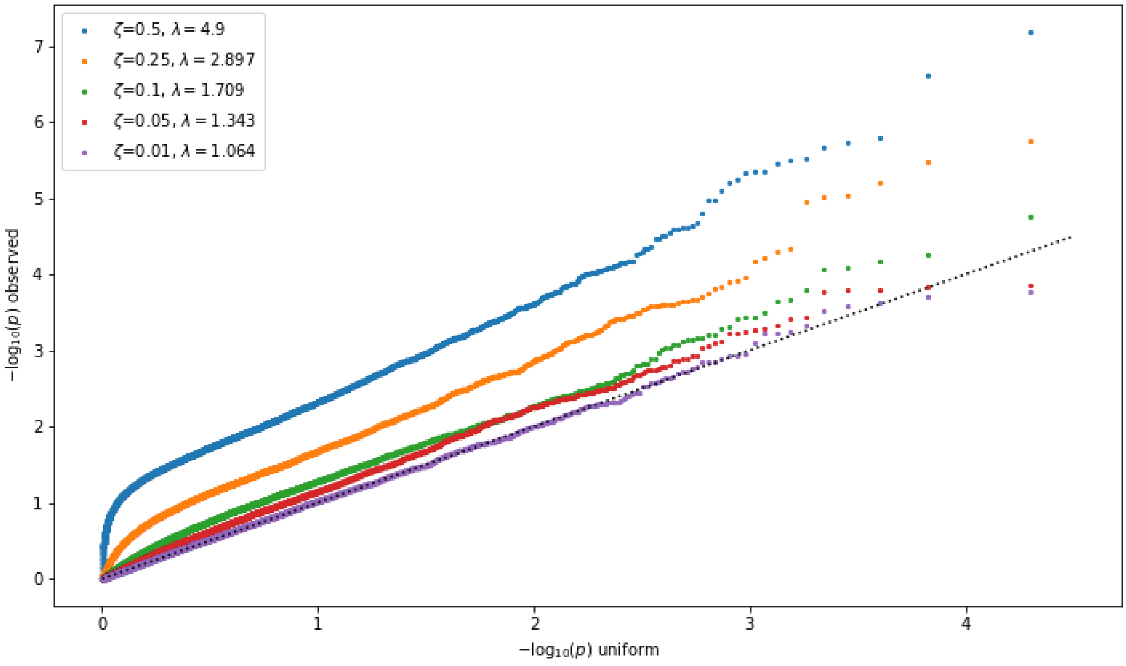


FIG. 3: QQ plot for simulation of the index (16) without applied correction for $\zeta$ for various values of $\zeta$. The control factor $\lambda$ is taken to be the median of observed $-\log_{10}$ transformed $p$-values divided by $-\log_{10}(0.5)$. An observed $\lambda < 1.1$ is commonly viewed as acceptable.

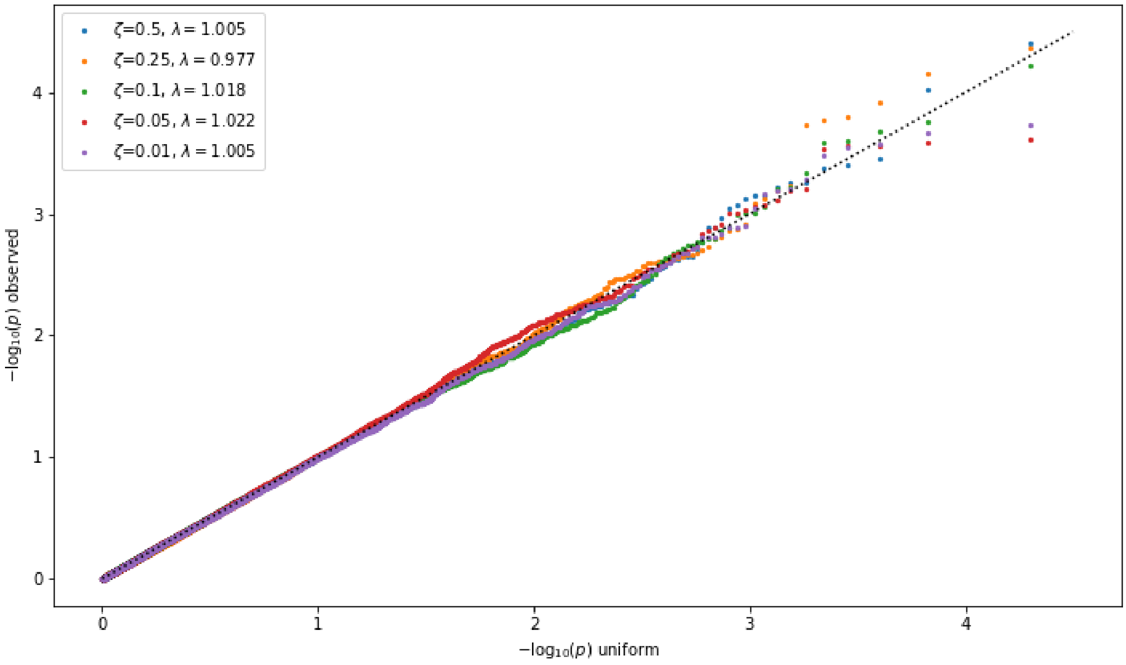


FIG. 4: QQ plot for simulation of the index (16) with applied correction for $\zeta$ for same values of $\zeta$ as in figure 3. $\lambda$ is defined as in figure 3. After applying the correction, the $p$-values are indeed well calibrated and $\lambda$ close to one.

with

$$t := \frac{\zeta - r}{\sqrt{1 + r^2 - 2\zeta r}} \, .$$

Similar to the main text, a consistency check can be performed against the Cauchy distribution. We know that for two normal distributed random variables $x \sim \mathcal{N}(0, \sigma_x^2)$ and $y \sim \mathcal{N}(0, \sigma_y^2)$ with correlation $\zeta$,

$$\frac{x}{y} \sim \mathcal{C}\left(\frac{\sigma_x}{\sigma_y}\zeta, \frac{\sigma_x}{\sigma_y}\sqrt{1-\zeta^2}\right) \, ,$$

with $\mathcal{C}$ the general Cauchy distribution. The corresponding cumulative distribution function for our case reads

$$F = \frac{1}{2} + \frac{1}{\pi} \arctan\left(\frac{r - \zeta}{\sqrt{1-\zeta^2}}\right) \, .$$

Explicit evaluation shows agreement with the $\chi^2$ based calculation via equation (22).

For partial sample overlap, the previous discussion for the coherence test applies one to one to the ratio test, and therefore the phenotypic correlation $\zeta$ has to be adjusted as well according to equation (19). As for the coherence, we calculate $p$-values for the index $R$ defined in equation (20) for 1000 random samples with and without correction factor $\zeta$. Corresponding QQ-plots are shown in figure 5 and 6. We observe that the correction indeed improves the calibration of $p$-values.

### V. SUPPLEMENTARY FIGURES FOR MAIN TEXT

- Figure 7: COVID-19 GWAS Manhattan plot.
- Figure 8: QQ plot for main text figure 5.
- Figure 9: QQ plot for different window sizes.
- Figure 10: SNP spectrum for *LZTFL1*.
- Figure 11: QQ plot for main text figure 8.
- Figure 12: SNP spectrum for *TRIM26*.
- Figure 13: QQ plot for main text figure 9.

### VI. SUPPLEMENTARY TABLES FOR MAIN TEXT

- Table II: Drug class codes.

| **Code** | **Drug class** |
|---|---|
| A02B | Drugs for peptic ulcer and gastro-oesophageal reflux disease |
| A10 | Drugs used in diabetes |
| B01A | Antithrombotic agents |
| C01D | Vasodilators used in cardiac diseasess |
| C02 | Antihypertensives |
| C03 | Diuretics |
| C07 | Beta blocking agents |
| C08 | Calcium channel blockers |
| C09 | Agents acting on the renin-angiotensin system |
| C10AA | HMG CoA reductase inhibitors |
| H03A | Thyroid preparations |
| L04 | Immunosuppressants |
| M01A | Anti-inflammatory and antirheumatic products, non-steroids |
| M05B | Drugs affecting bone structure and mineralization |
| N02A | Opioids |
| N02BA | Salicylic acid and derivatives |
| N02BE | Anilides |
| N02C | Antimigraine preparations |
| N06A | Antidepressants |
| R03A | Adrenergics, inhalants |
| R03BA | Glucocorticoids |
| R06A | Antihistamines for systemic use |
| S01E | Antiglaucoma preparations and miotics |

TABLE II: Drug codes and corresponding classes for the drug GWASs of [7] used in the main text.

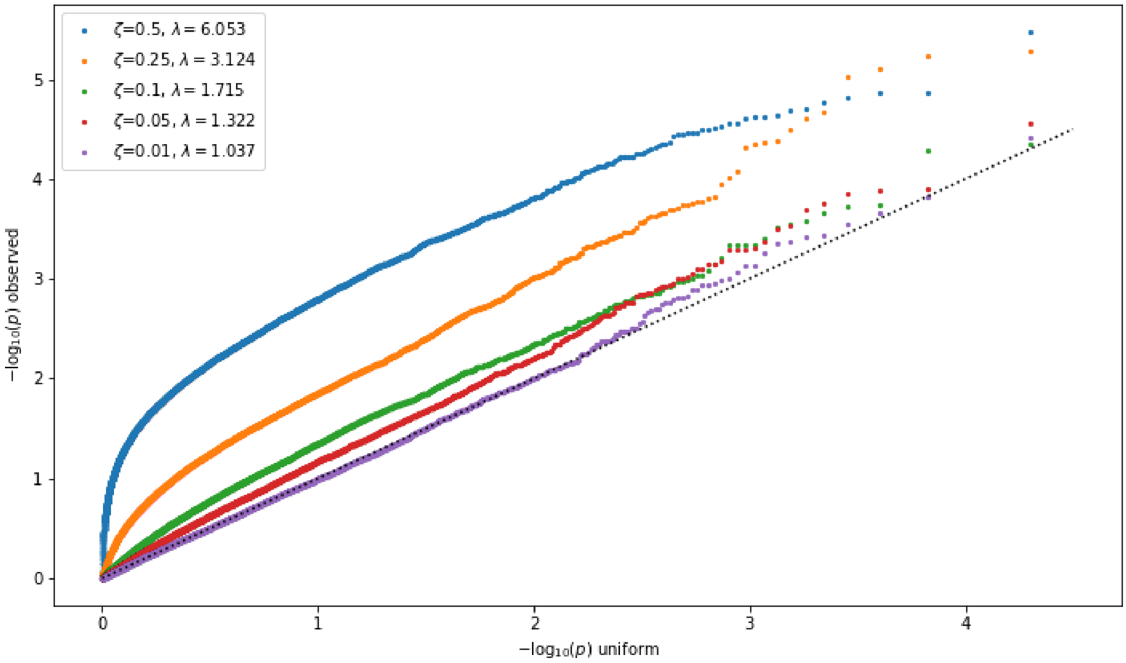


FIG. 5: QQ plot for simulation of the index (20) without applied correction for $\zeta$ for various values of $\zeta$. $\lambda$ is defined as in figure 3.

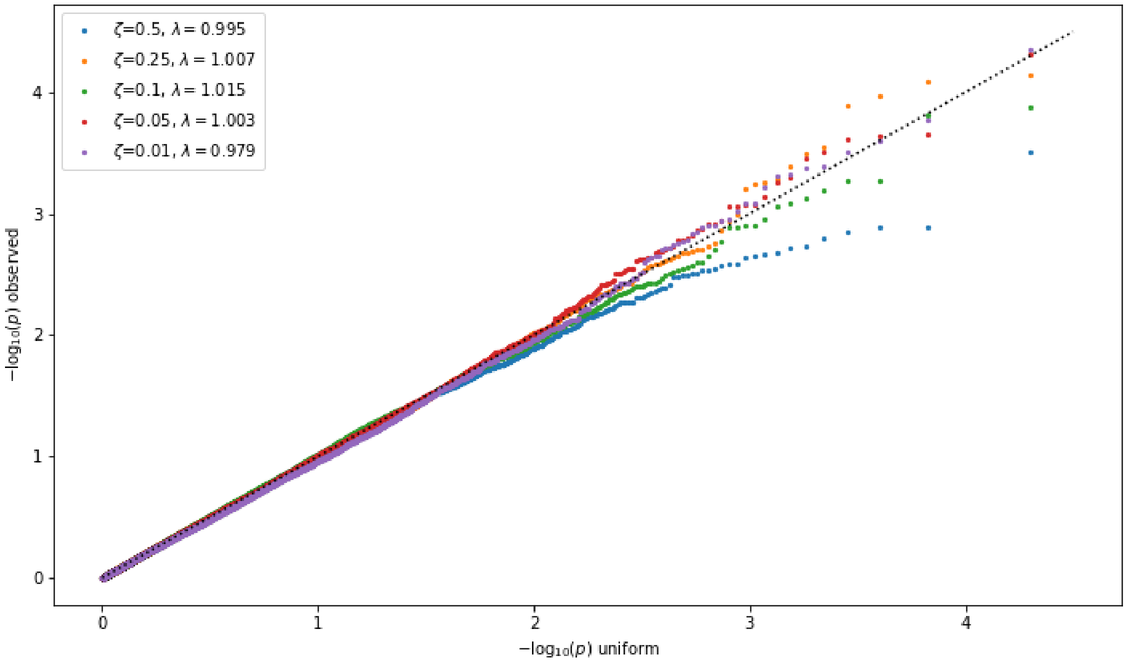


FIG. 6: QQ plot for simulation of the index (20) with applied correction for $\zeta$ for same values of $\zeta$ as in figure 5. $\lambda$ is defined as in figure 3. After applying the correction, the $p$-value calibration improved significantly and $\lambda$ is close to one.

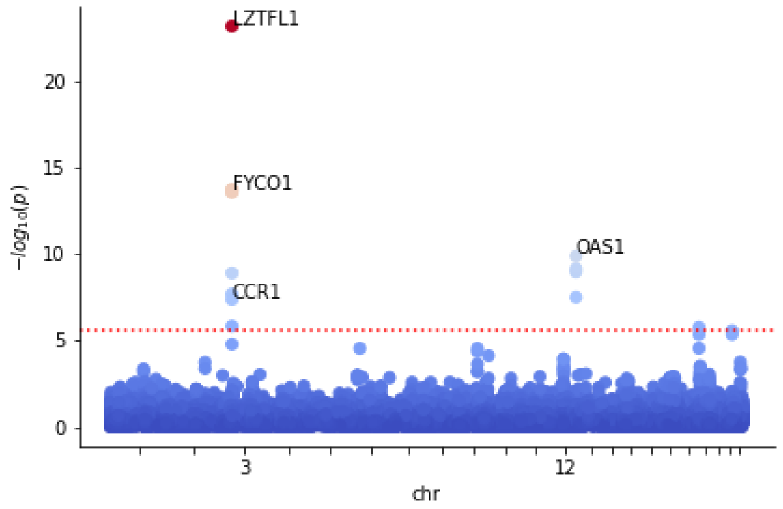


FIG. 7: Manhattan plot showing the strong gene enrichment on chromosomes 3 and 12 for the severe COVID-19 GWAS. The Bonferroni significance threshold (red dotted line) is taken to be $2.7 \times 10^{-6}$ (0.05 divided by 18453, the number of tested genes). Only a selection of significant genes is labeled.

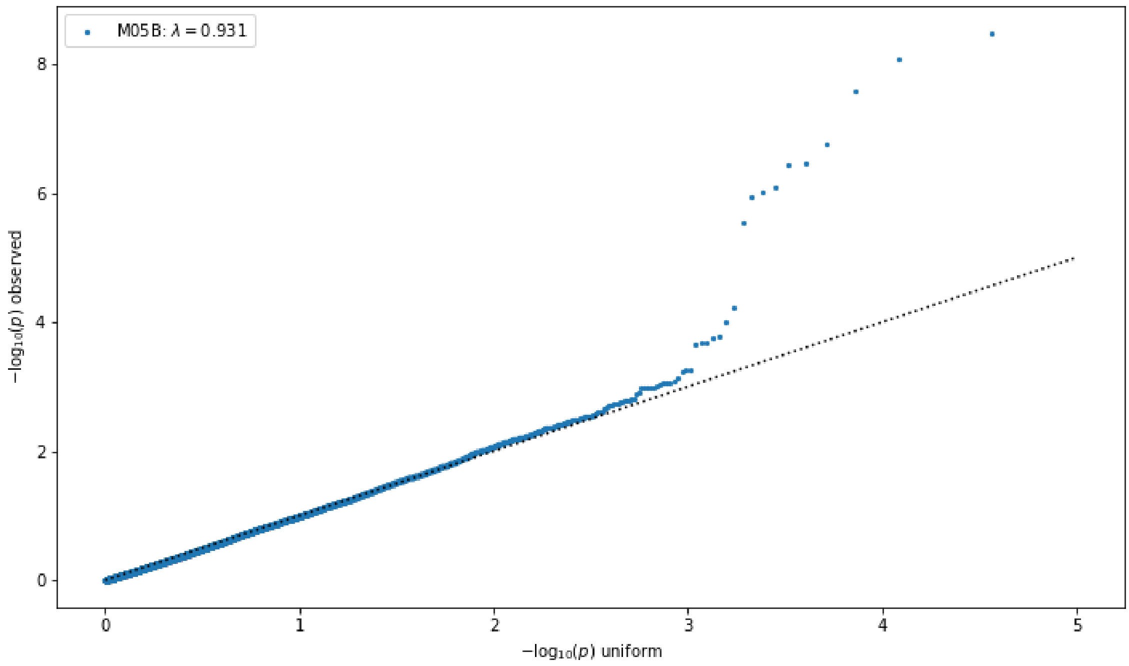


FIG. 8: QQ plot for coherent cross-scoring of COVID-19 against drug class M05B. Each data point corresponds to a gene. The European sub-population of the 1K Genome project has been used as a reference panel. $\lambda$ is defined as in figure **??**.

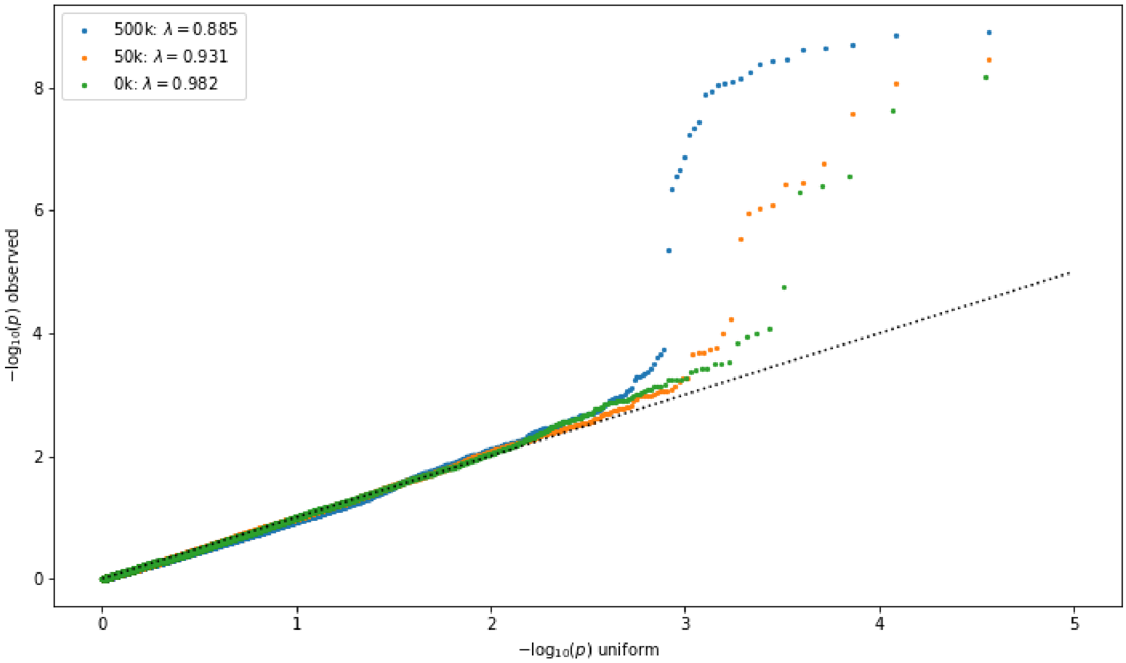


FIG. 9: QQ plot for coherent cross-scoring of COVID-19 against drug class M05B for different gene window sizes. Each data point corresponds to a gene. Reference panel and $\lambda$ as for figure 8.

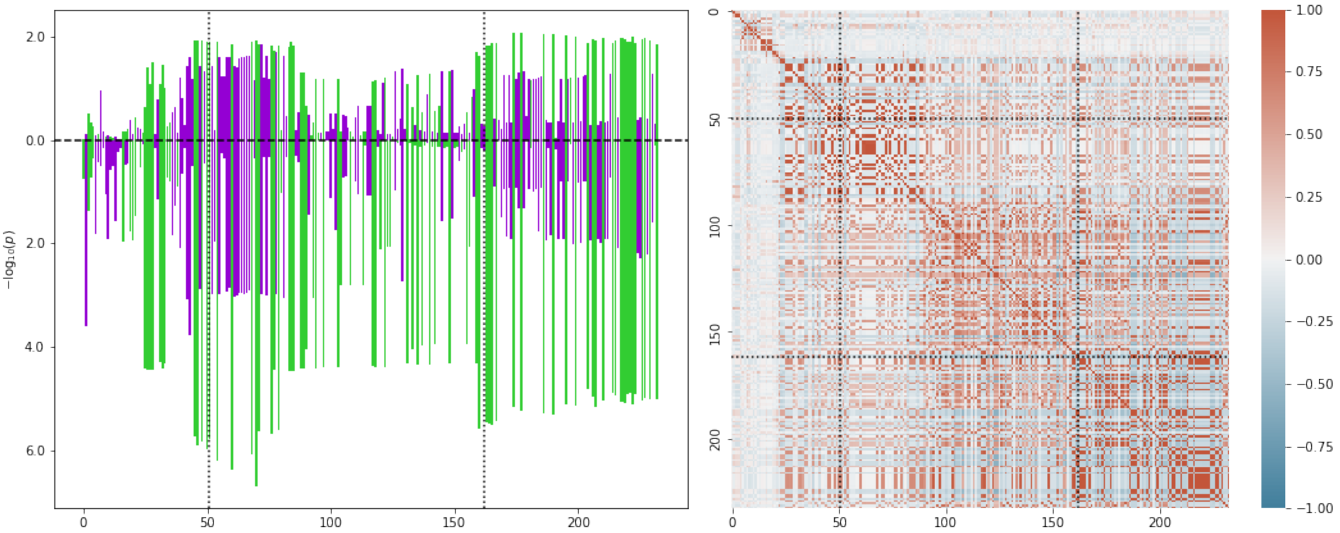


FIG. 10: Left: SNP $p$-values after rank transform for the *LZTFL1* gene. The $x$-axis is numbered according to the $i$th SNP in the gene window ordered by increasing position. The dotted black lines indicate the transcription start and end positions (first and last SNP). Up bars correspond to M05B and down bars to severe COVID-19. Green indicates positive and violet negative association with the trait. Right: SNP-SNP correlation matrix inferred from the 1KG reference panel. Note that the gene contains at least three sizable LD blocks.

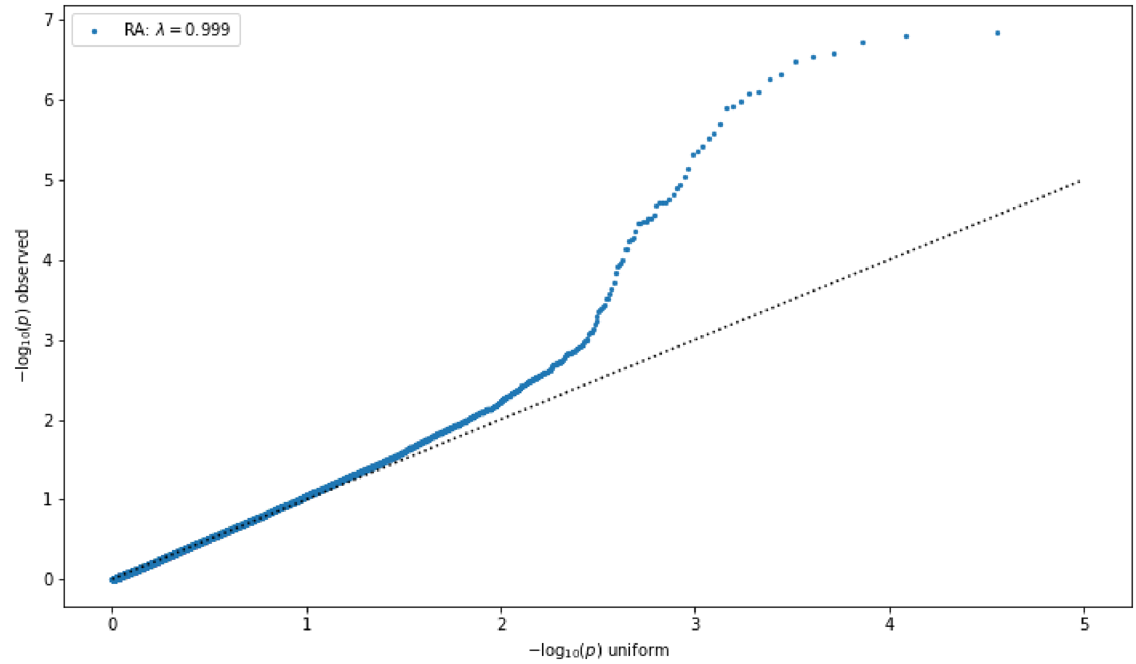


FIG. 11: QQ plot for anti-coherent cross-scoring of COVID-19 against rheumatoid arthritis. Caption otherwise as in figure 8.

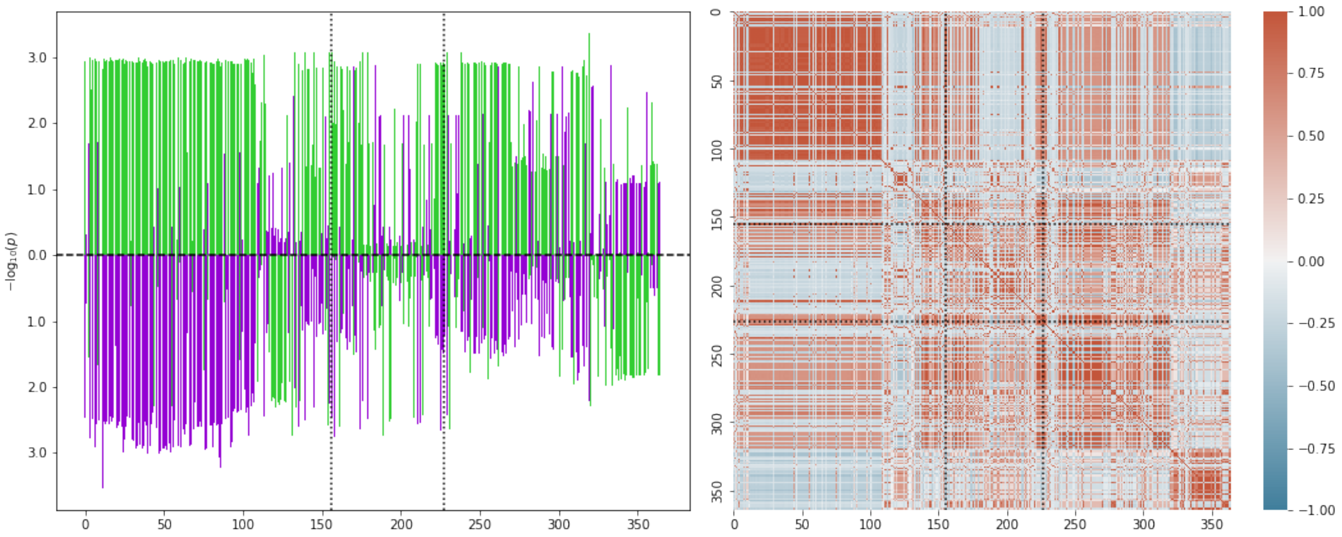


FIG. 12: Left: SNP $p$-values after rank transform for the *TRIM26* gene under medication L04. Annotation as in figure 10, but up bars corresponding to L04. Right: SNP-SNP correlation matrix inferred from the 1KG reference panel.

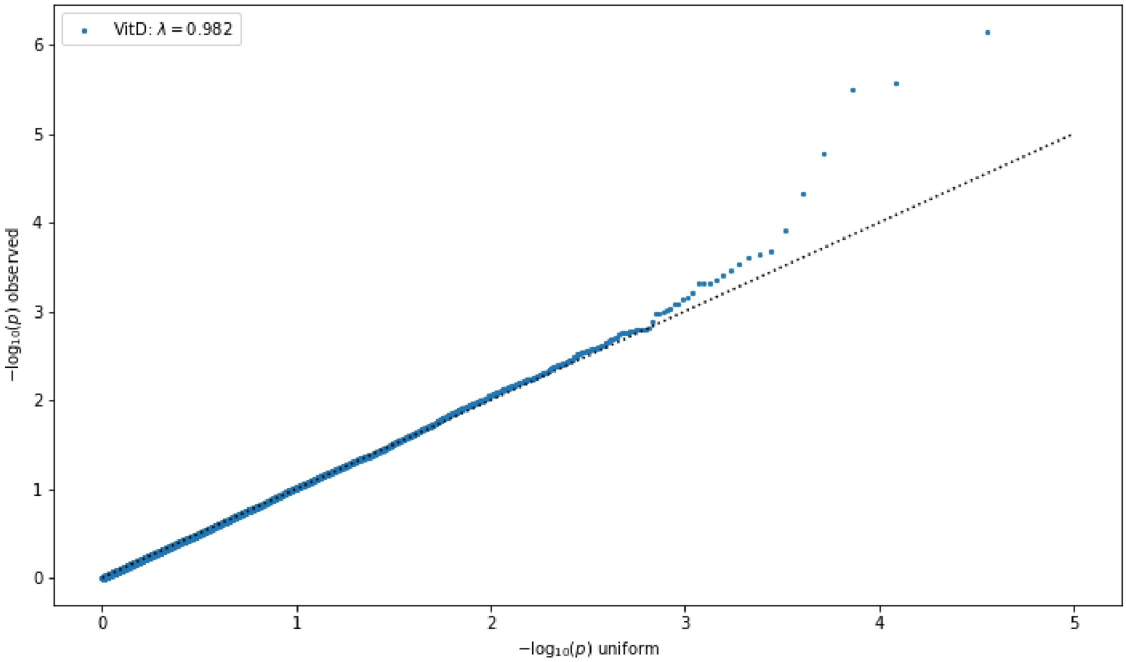


FIG. 13: QQ plot for ratio scoring Vitamin D concentration against severe COVID-19 in the coherent case, with ratio denominator given by COVID-19. Caption otherwise as in figure 8.

[8] Lamparter D, Marbach D, Rueedi R, Kutalik Z, and Bergmann S. "Fast and rigorous computation of gene and pathway scores from SNP-based summary statistics." PLoS Computational Biology 12, e1004714, 2016

[9] Bulik-Sullivan B, Finucane HK, Anttila V, et al. "An atlas of genetic correlations across human diseases and traits." Nat Genet. 2015;47(11):1236-1241. doi.org/10.1038/ng.3406
